# Fractional flow reserve versus angiography for revascularization in patients with coronary artery disease: A systematic review and meta-analysis of randomized controlled trials

**DOI:** 10.1101/2023.08.18.23294291

**Authors:** Sunny Goel, Sharon Slomovich, Sami Edris, Gal Rubinstein, Chirag Agarwal, Won Jun Park, Daniel Zinkovsky, Amit Hooda, Parasuram Melarcode Krishnamoorthy, Umesh K. Gidwani, Samin K. Sharma, Annapoorna Kini

## Abstract

**Background:** Fractional flow reserve (FFR) is routinely used to assess the ischemic potential of a coronary artery lesion. However, recently published randomized control trials have questioned the advantage of FFR over angiography to guide revascularization. Whether FFR guided revascularization provides clinical benefit over angiography remains unclear.

**Methods:** We performed a meta-analysis in patients with stable coronary artery disease (CAD), acute coronary syndrome (ACS), multivessel or single vessel CAD undergoing revascularization comparing FFR versus angiography to guide revascularization. Randomized control trials comparing FFR versus angiography guided revascularization were searched through PubMed, Cumulative Index to Nursing and Allied Health Literature, Cochrane Central, Scopus, Google Scholar, and Web of Science databases. The primary endpoints included cardiovascular mortality, repeat revascularization, myocardial infarction, major adverse cardiac events, stroke or transient ischemic attack and target lesion revascularization. We also evaluated the procedural outcomes including the average number of stents used between the two groups, procedure time and contrast volume used. Event rates were compared using a forest plot of odds ratios using a random-effects model assuming interstudy heterogeneity.

**Results:** The meta-analysis included 13 trials in which 7415 patients met the eligibility criteria. There was no significant difference between the FFR versus angiography guided revascularization groups across all clinical measures including all-cause mortality (OR = 1.06, 95% CI = 0.74-1.53, P = 0.74, I2= 27%), cardiovascular mortality (OR = 0.81, 95% CI = 0.43-1.52, P = 0.51, I^2^= 44%), repeat revascularization (OR = 1.02, 95% CI = 0.83-1.26, P = 0.83, I^2^= 17%), myocardial infarction (OR = 0.92, 95% CI = 0.69-1.21, P = 0.54, I^2^= 36%), major adverse cardiac event (OR = 0.82, 95% CI = 0.62-1.08, P = 0.15, I^2^= 41%), stroke or transient ischemic attack (OR = 1.49, 95% CI = 0.87-2.55, P = 0.15, I^2^= 0%) and target lesion revascularization (OR = 0.86, 95% CI = 0.44-1.69, P = 0.67, I^2^= 0%). A sensitivity analysis was performed for studies that included patients exclusively with an ACS and studies that used FFR coronary artery bypass grafting (CABG) as a revascularization strategy. There was no difference in any of the clinical outcomes between the two groups in the sensitivity analysis. In terms of procedural outcomes, the average number of stents used was lower in the FFR group as compared to the angiography group, mean difference (MD) of −0.79 (95% CI = − 1.10, − 0.48), P < 0.00001) with no difference in procedure time or contrast volume used.

**Conclusion:** This meta-analysis suggests that FFR when used in conjunction with angiography prevents unnecessary PCI without any difference in clinical outcomes between the two groups.

## INTRODUCTION

In patients with coronary artery disease (CAD), localizing lesions responsible for reversible ischemia can be challenging. Identifying these physiologically significant lesions by coronary angiography, particularly those of indeterminate significance (40-70% diameter narrowing) is subject to limitations due to inter-operator variability and inaccuracies in visual interpretation^1–4^. Fractional flow reserve (FFR) is routinely used to estimate the functional impact of these intermediate coronary stenosis and to guide percutaneous intervention in everyday practice^3,5^.

Fractional flow reserve is a diagnostic technique that measures pressure differences proximal and distal to a stenotic coronary lesion at maximal flow, indicating the degree of stenosis. FFR is utilized during coronary angiography to guide revascularization by assessing the ischemic potential of a coronary artery lesion^4,6,7^. An FFR index of ≤0.80 identifies a coronary lesion causing ischemia with >90% accuracy. Clinical trials have demonstrated that FFR-guided angiography can prevent unnecessary percutaneous intervention and improve clinical outcomes^7–11^.

The advantages of FFR over angiography to guide revascularization have been well established and FFR has remained the clinical standard in guiding clinical decision-making for lesions with questionable hemodynamic impact^3,10,12^. However, recently published randomized controlled trials have questioned the clinical benefits of using FFR over angiography to guide coronary artery revascularization in patients with multi-vessel CAD, especially in patients with acute coronary syndromes ^13–16^. We, therefore, conducted a meta-analysis of all randomized control trials to compare the clinical outcomes in patients with stable CAD, acute coronary syndrome, single vessel CAD and multivessel disease who underwent FFR versus angiography-guided revascularization either with PCI or CABG.

## MATERIALS AND METHODS

### Study Design

A systematic review of the literature was performed according to the Preferred Reporting Items for Systematic Reviews and Meta-Analyses (PRISMA) statement^17^.

### Data sources and search strategy

We conducted a systematic search of electronic databases including PubMed, Cumulative Index to Nursing and Allied Health Literature (CINAHL), Cochrane Central, Scopus, Google Scholar, and Web of Science databases for all randomized control trials (RCT) that compared FFR versus angiography for patients with CAD. All relevant combinations of keywords relating to FFR and angiography for CAD were searched: “fractional flow reserve”, “angiography”, “revascularization,” “multi-vessel coronary artery disease” and “randomized control trial.” The search was conducted from the inception of these databases to December 31st, 2022. No language or age restrictions were applied. Pertinent trials were also searched on http://www.clinicaltrials.gov and in the proceedings of major international cardiology meetings (American College of Cardiology, American Heart Association, European Society of Cardiology, and Transcatheter Cardiovascular Therapeutics [TCT]). We also manually searched the references of these articles to find additional relevant articles. Two independent reviewers SG and SS conducted the search. This meta-analysis was registered at PROSPERO (CRD42022301144). This study was exempted from Institutional Review Board approval, as it was a study-level meta-analysis.

### Study Selection

We included in our meta-analysis all studies that met the following criteria: (1) randomized control trial (2) study on human subjects with participants of any age (3) study reporting clinical outcomes between FFR versus angiography guided revascularization either with PCI or CABG (4) full-length article in a peer-reviewed journal.

In addition, we also included the FAME 3 trial, one of the largest clinical trials evaluating the role of physiological assessment of coronary artery lesions to provide additional strength to our analysis. It compared FFR guided percutaneous intervention (PCI) versus angiography guided CABG in patients with 3 vessel CAD^18^. Although PCI and CABG are different means of revascularization, we wanted to evaluate the totality of the evidence. CABG is routinely used to revascularize patients with multivessel CAD and FFR is employed during the pre-CABG diagnostic angiography to determine hemodynamic significance of intermediate coronary artery lesions. Separate sensitivity analyses were performed to exclude the FAME 3 trial (Figure S1), studies with ACS patients exclusively (Figure S2) and the studies that used FFR guided CABG as a revascularization strategy (Figure S3). The longest follow-up data available for the included studies was used in the final analysis.

Several trials that randomized patients to culprit versus non-culprit lesions, with subsequent FFR versus angiography guided revascularization, such as the COMPLETE and CVLPRIT trails, were excluded from our analysis as they did not strictly randomize patients to FFR versus angiography^19,20^. FFR was left at the operator’s discretion in these trials. The recently published RIPCORD-2 trial, and FRAME AMI (acute myocardial infarction) trials are also included in our analysis^16,21^. For our analysis, the events rates were calculated only for the patients with angiographic obstructive coronary disease in accordance with previous trials^16^.

### Data Extraction

Reviewers (SG and SS) independently screened the titles and abstracts that met the inclusion criteria. Discrepancies between reviewers were discussed until a consensus was reached. Using the above-mentioned selection criteria, these two reviewers independently determined which articles were to be included and excluded, and the data from the relevant articles were extracted using predefined extraction forms. Any disagreements in data extraction were discussed until a consensus was reached. Bibliographies of relevant publications were hand-searched to attempt complete inclusion of all studies of interest.

### Study endpoints

The clinical endpoints included were all-cause mortality, cardiovascular mortality, repeat revascularization including target vessel revascularization, myocardial infarction (MI), major adverse cardiac event (MACE), stroke or transient ischemic attack (TIA), and target lesion revascularization (TLR). The definition of outcomes as described in the trials are defined in Table S1. The procedural outcomes evaluated were the average number of stents used, total contrast volume and procedure time.

### Data analysis

To analyze the data, we used Review Manager Software (Rev Man, version 5.3). Analysis was performed on an intention-to-treat basis. Data were summarized across treatment arms using the Mantel– Haenszel odds ratio (OR) using a random-effects model. Heterogeneity between studies was assessed using the Cochrane Q test and I^2^ statistics, which denotes the percentage of total variation across studies that is a result of heterogeneity rather than chance. Heterogeneity was considered significant if the p-value was <0.05. Sensitivity analyses were performed as described in the study selection.

### Risk of bias assessment

The risk of the bias for the included trials was evaluated using the Cochrane risk assessment tool for bias, which comprises 7 criteria: random sequence generation, allocation concealment, blinding of participants and personnel, blinding of outcome assessment, incomplete outcome data, selective reporting, and other sources of bias. Based on the fulfillment of these criteria, studies were classified as low risk, unclear risk, or high risk for bias. Publication bias was evaluated using the Egger test and funnel plots were created (Figure S4).

The datasets generated during and/or analyzed during the current study are available from the corresponding author on reasonable request.

## RESULTS

Figure 1 shows the Preferred Reporting Items for Systematic Reviews and Meta-Analyses (PRISMA) flow diagram describing the search strategy. The initial search yielded 2,163 abstracts of which 1,849 were excluded based on the title and reading of the abstract. Six hundred and seventy-five articles were reviewed out of which 361 were excluded as they were either non-randomized studies or had no comparison group.

Table 1 outlines the included studies for the final analysis. A total of thirteen RCTs met the inclusion criteria. The weighted mean follow-up period was 22 months (1.8 years) with a total of 7415 patients (FFR group = 3719, angiography group = 3714).

**Table 1.**
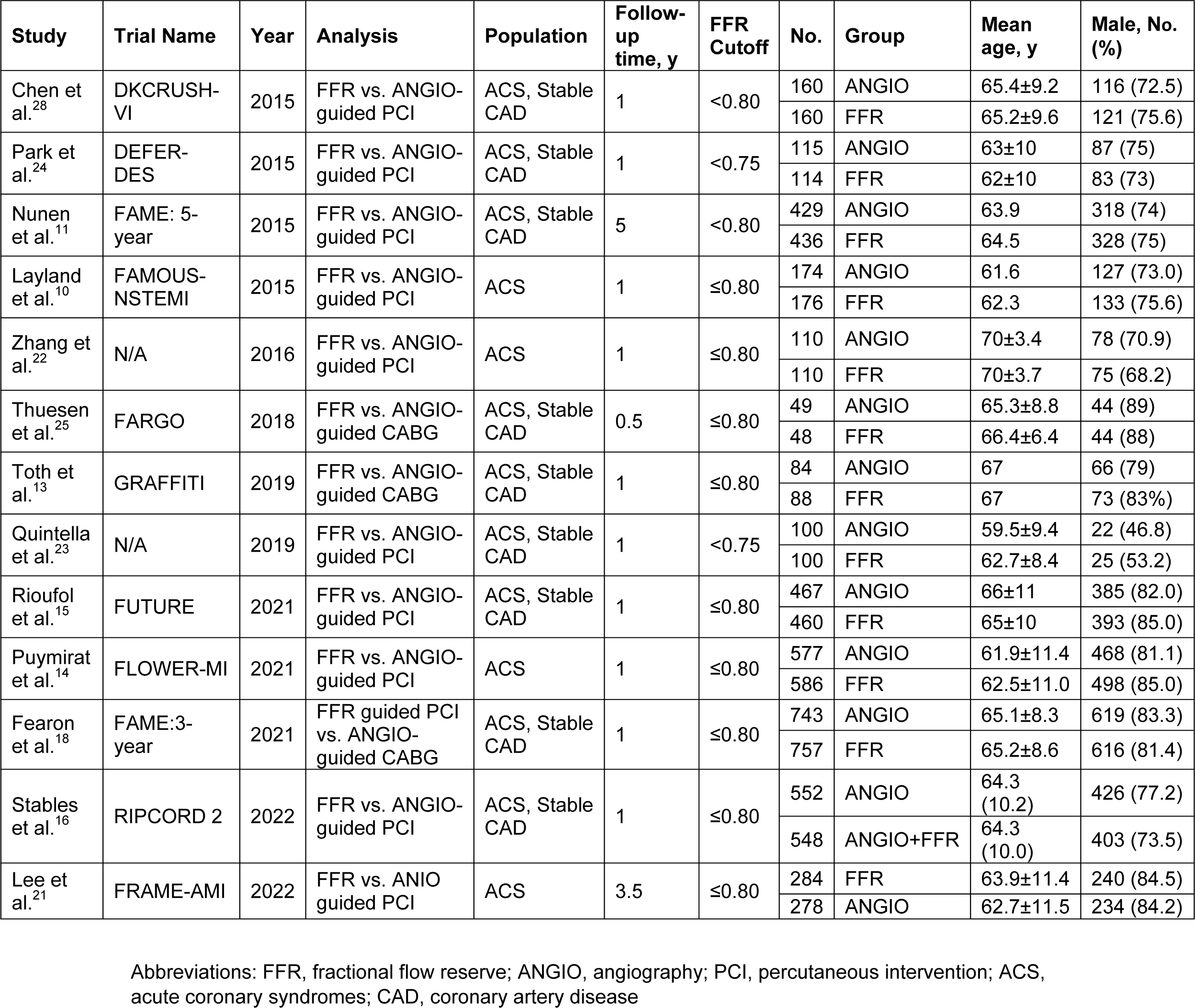
Baseline Characteristics.

The baseline characteristics were similar between both groups (Table 1). The mean age of patients in the angiography group was 64.4 ± 2.8 years, whereas it was 64.8 ± 2.2 years in the FFR group. Males comprised 77.4% of patients in the angiography group compared to 77.9% of the patients in the FFR group. A history of diabetes mellitus was present in 23.8% of the patients in the angiography group and 24.4% of the patients in the FFR group. The prevalence of hypertension was 59.7% and 58.7% in the angiography and FFR groups respectively. Of the patients in the angiography group, 58.3 % and 57.3% of patients in the FFR group had a history of hyperlipidemia. In the angiography group, 33.9% of patients were current smokers compared to 33.0% of those in the FFR group. Previous MI was noted in 18.0% of patients in the angiography group and 21.6% of patients in the FFR group (Table 2).

**Table 2.** Baseline comorbidities.

| Study | Group | Diabetes Mellitus, No. (%) | Hypertension, No. (%) | Hyperlipidemia, No. (%) | Current smoker, No. (%) | Previous Myocardial Infarction, No. (%) |
| --- | --- | --- | --- | --- | --- | --- |
| Chen et al.<sup>28</sup> | ANGIO | 43 (26.9) | 106 (68.3) | 32 (20.0) | 64 (40.0) | 19 (11.9) |
|  | FFR | 48 (30.0) | 116 (72.5) | 27 (16.9) | 66 (41.3) | 12 (7.5) |
| Park et al.<sup>24</sup> | ANGIO | 39 (34) | 65 (57) | 78 (68) | 38 (33) | 20 (17) |
|  | FFR | 30 (26) | 73 (64) | 80 (70) | 30 (26) | 22 (19) |
| Nunen et al.<sup>11</sup> | ANGIO | 107 (25) | 277 (65) | 316 (74) | 130 (30) | 155 (36) |
|  | FFR | 98 (22) | 259 (59) | 307 (70) | 111 (25) | 154 (35) |
| Layland et al.<sup>10</sup> | ANGIO | 26 (14.9) | 81 (46.6) | 56 (32.2) | 71 (40.8) | 24 (13.8) |
|  | FFR | 26 (14.8) | 78 (44.3) | 71 (40.3) | 72 (40.9) | 22 (12.5) |
| Zhang et al.<sup>22</sup> | ANGIO | 36 (32.7) | 83 (75.5) | 93 (84.5) | 31 (28.2) | 23 (20.9) |
|  | FFR | 40 (36.4) | 81 (73.6) | 90 (81.8) | 29 (26.4) | 24 (21.8) |
| Thuesen et al.<sup>25</sup> | ANGIO | 11 (23) | 33 (67) | 36 (75) | 8 (17) | 10 (21) |
|  | FFR | 11 (22) | 33 (67) | 42 (86) | 13 (27) | 14 (29) |
| Toth et al.<sup>13</sup> | ANGIO | 33 (40) | 59 (70) | 66 (79) | 35 (42) | 5 (6) |
|  | FFR | 31 (35) | 68 (77) | 70 (80) | 47 (53) | 15 (17) |
| Quintella et al.<sup>23</sup> | ANGIO | 12 (50.0) | 26 (50.9) | 26 (52.0) | 9 (47.4) | 7 (46.7) |
|  | FFR | 12 (50.0) | 25 (49.0) | 24 (42.0) | 10 (52.6) | 8 (53.3) |
| Rioufol et al.<sup>15</sup> | ANGIO | 147 (32.0) | 283 (61.0) | 286 (61.0) | 118 (26.0) | 100 (21.0) |
|  | FFR | 143 (31.0) | 265 (58.0) | 275 (60.0) | 109 (24.0) | 90 (20.0) |
| Puymirat et al.<sup>14</sup> | ANGIO | 82(14.2) | 262 (45.4) | 237 (41.1) | 210 (36.4) | 31 (5.4) |
|  | FFR | 107(18.3) | 253 (43.2) | 232 (39.6) | 235 (40.1) | 45 (7.7) |
| Fearon et al.<sup>18</sup> | ANGIO | 214 (28.8) | 556 (75.0) | 531 (71.7) | 136 (18.4) | 248 (33.5) |
|  | FFR | 214 (28.3) | 538 (71.2) | 521 (68.9) | 145 (19.2) | 252 (33.3) |
| Stables et al.<sup>16</sup> | ANGIO | 97 (17.6) | 294 (53.5) | 317 (57.6) | 356 (65) | 129 (23.4) |
|  | ANGIO+FFR | 113 (20.6) | 315 (57.6) | 315 (57.5) | 316 (58.5) | 117 (21.4) |
| Lee et al.<sup>21</sup> | ANGIO | 86 (30.9) | 152 (54.7) | 107 (38.5) | 105 (37.8) | 7 (2.5) |
|  | FFR | 97 (34.2) | 151 (53.2) | 121 (42.6) | 91 (32.0) | 7 (2.5) |

Abbreviations: FFR, fractional flow reserve; ANGIO, angiography

## STUDY OUTCOMES

There was no significant difference between FFR versus angiography across all clinical outcomes; all-cause mortality (OR = 1.06, 95% CI = 0.74-1.53, P = 0.74, I2= 27%) [Figure 2], cardiac mortality (OR = 0.81, 95% CI = 0.43-1.52, P = 0.51, I2= 44%) [Figure 2], repeat revascularization (OR = 1.02, 95% CI = 0.83-1.26, P = 0.83, I2= 17%) [Figure 2], target lesion revascularization (OR = 0.86, 95% CI = 0.44-1.69, P = 0.67, I2= 0%) [Figure 2], major adverse cardiac events [MACE] (OR = 0.82, 95% CI = 0.62-1.08, P = 0.15, I2= 41%) [Figure 2], myocardial infarction (OR = 0.92, 95% CI = 0.69-1.21, P = 0.54, I2= 36%) [Figure 2], incidence of Stroke or TIA (OR = 1.49, 95% CI = 0.87-2.55, P = 0.15, I2= 0%) [Figure 2]. In terms of procedural outcomes, the average number of stents used was lower in the FFR group as compared to the angiography group, MD of − 0.79 (95% CI = − 1.10, − 0.48), P < 0.00001, with no difference in procedure time or contrast volume used [Figure 3].

**Figure 1.**
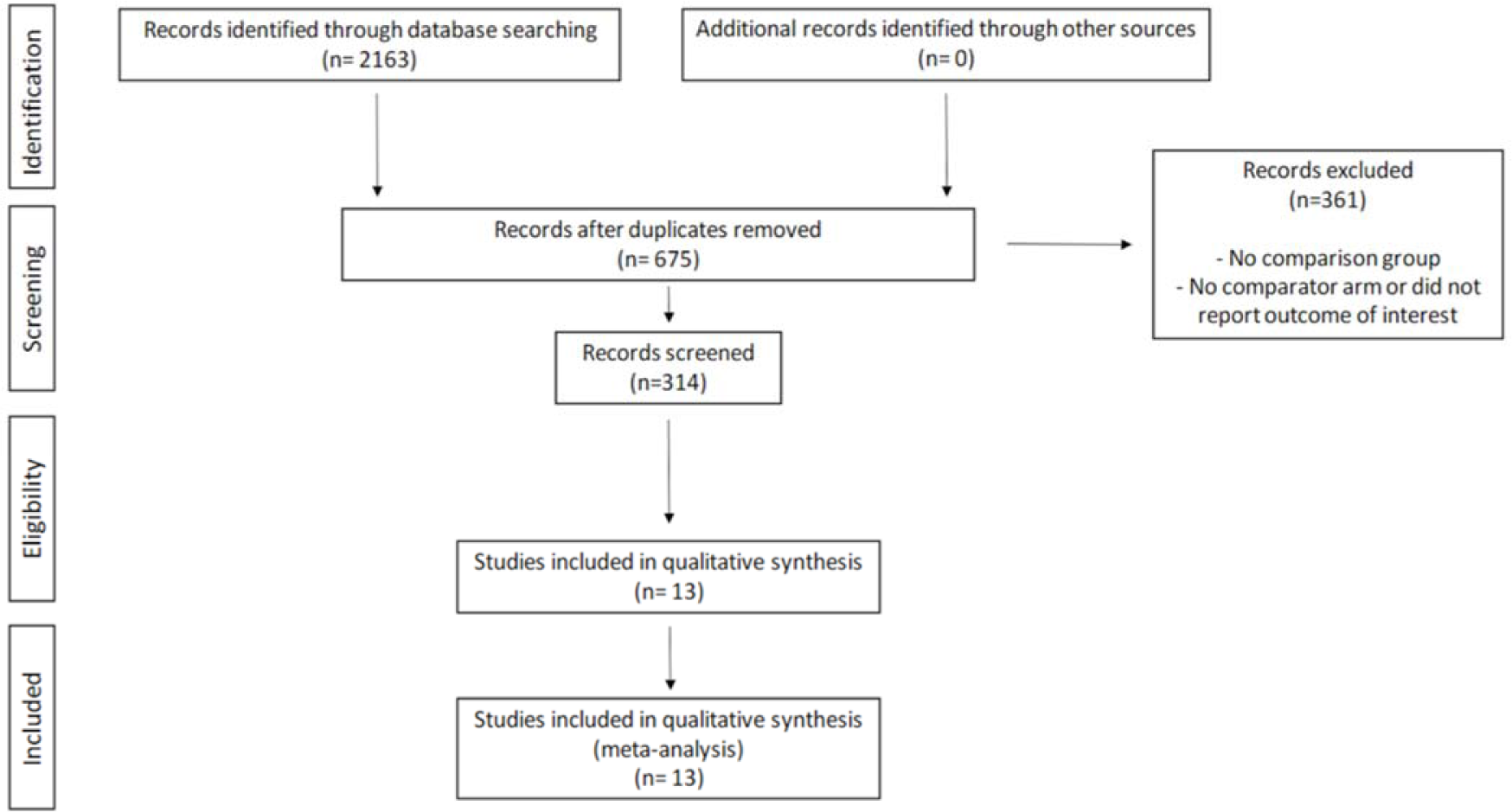
Preferred Reporting Items for Systemic Reviews and Meta-Analysis (PRISMA) flow diagram.

**Figure 2.**
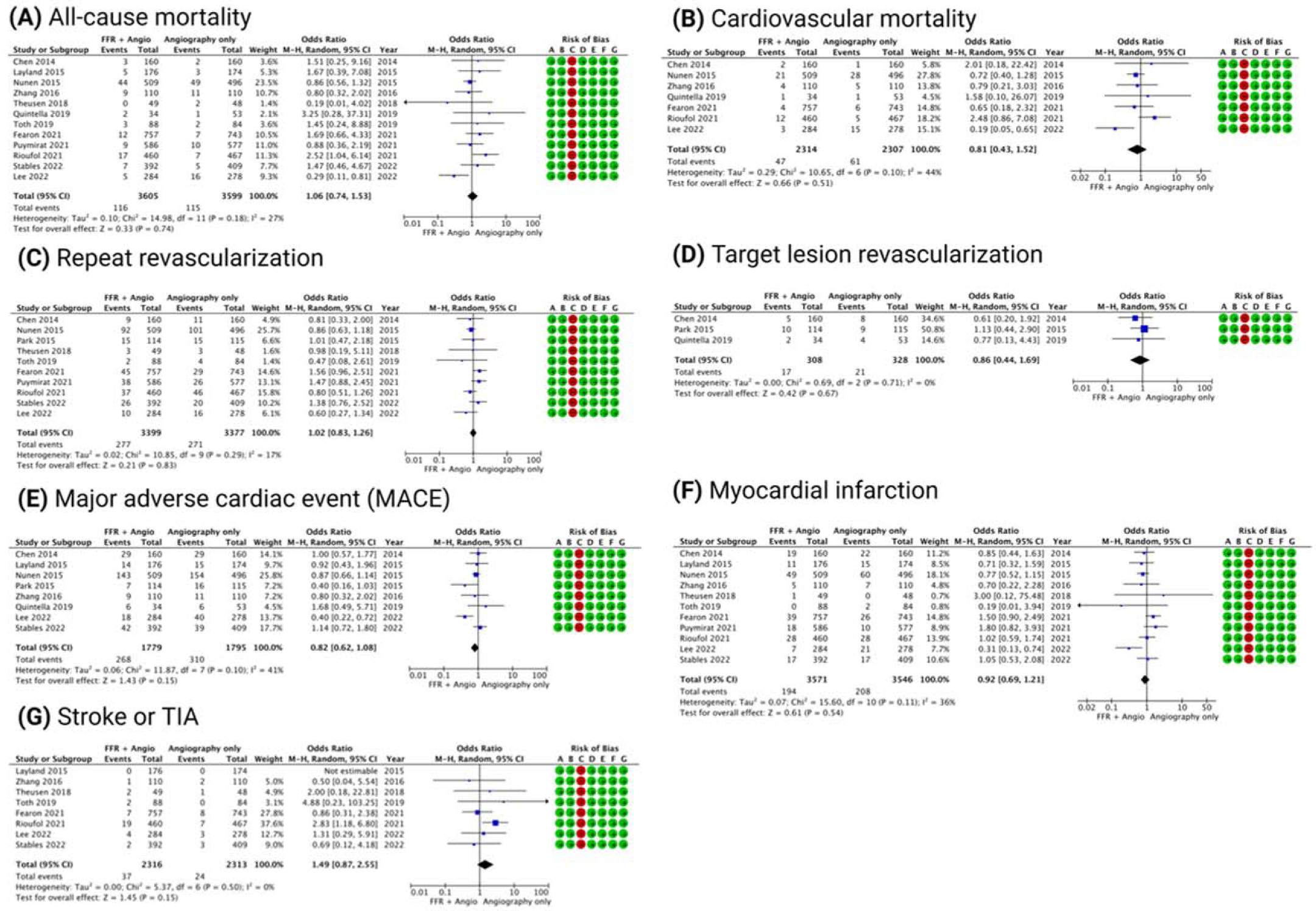
(A) Forest plot showing all-cause mortality comparing FFR versus angiography (B) Forest plot showing cardiac mortality comparing FFR versus angiography (C) Forest plot showing repeat revascularization comparing FFR versus angiography (D) Forest plot showing target lesion revascularization (TLR) comparing FFR versus angiography (E) Forest plot showing MACE comparing FFR versus angiography (F) Forest plot showing myocardial infarction comparing FFR versus angiography (G) Forest plot showing stroke or TIA comparing FFR versus angiography.

**Figure 3.**
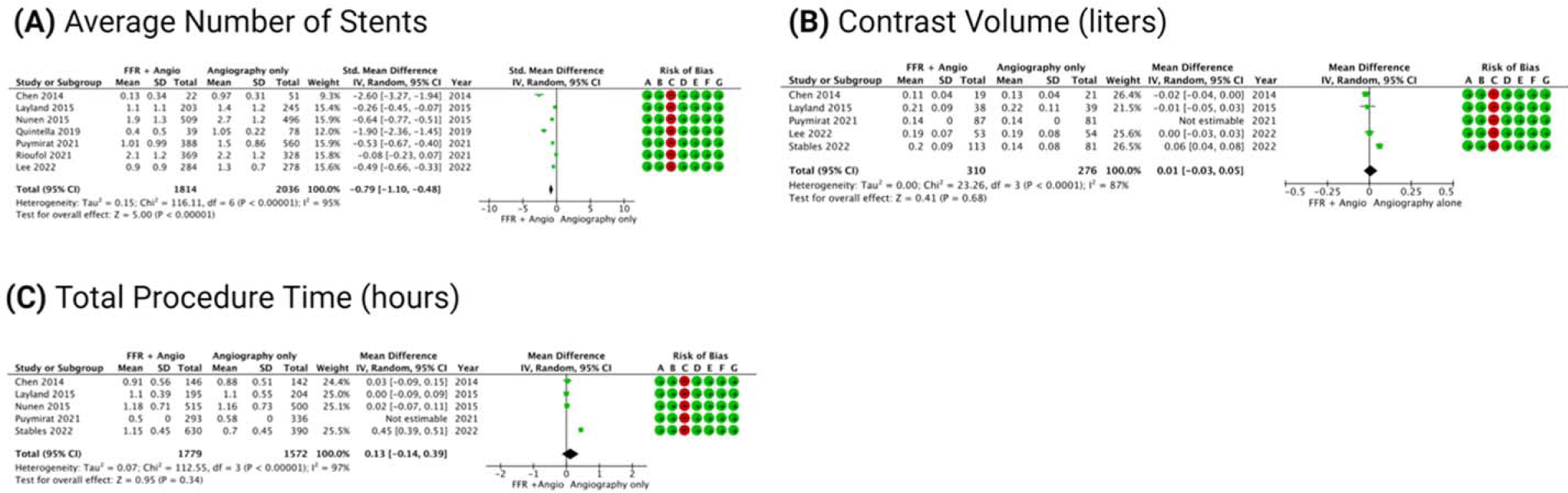
(A) Forest plot showing the average number of stents (B) Forest plot showing contrast volume (C) Forest plot showing total procedural time

A sensitivity analysis was performed for studies with ACS patients exclusively and the studies that used FFR guided CABG as a revascularization strategy. There was no statistically significant difference between the two groups (Figure S2 and S3).

## DISCUSSION

The main finding of our analysis is that there is no significant difference in clinical outcomes (all-cause mortality, cardiovascular mortality, repeat revascularization, MI, MACE, Stroke/TIA, TLR) in patients undergoing FFR versus conventional angiography guided revascularization at the mean follow-up period of 1.8 years. FFR does however reduce the average number of stents used, with no difference in the contrast volume used and procedure times.

FFR is currently utilized as the gold standard for determining the hemodynamic influence of an intermediate coronary lesion. FFR-guided PCI has been shown to reduce unnecessary revascularization and improve clinical outcomes in both single and multivessel CAD^3,11,12^. The FAME trial was one of the earliest multicenter, randomized control trials to demonstrate favorable clinical outcomes of FFR as compared to angiography to guide PCI in multivessel CAD^9^. In this cohort, there were a significantly higher number of stents placed in the angiography-guided group as compared to the FFR-guided group with a significant decrease in major adverse cardiac events in the FFR group in the first two years^11^. Several RCTs followed with encouraging results in patients with both acute coronary syndromes (ACS) and stable CAD^10,11,22–24^. However, recent studies have emerged questioning the role of FFR versus angiography to guide revascularization. In the FUTURE trial, there was no evidence to support that the FFR-guided treatment strategy reduced the risk of ischemic cardiovascular events or death at 1-year-follow-up^15^. Of note, the Data Safety and Monitoring Board of the FUTURE trial recommended early discontinuation of the trial for safety concerns as the all-cause mortality was increased in the FFR group as compared to the control group (4.3% versus 1.8%; P = 0.038). However, this difference in death was not statistically significant after the collection of all data and the final analysis (P = 0.06). Similarly, the recently published RIPCORD-2 trial showed no difference in clinical outcomes between the strategy of systematic FFR assessment compared with angiography^16^.

In the FAMOUS-NSTEMI trial, there were no statistically significant differences in clinical outcomes between the FFR guided revascularization versus routine angiography guided revascularization of non-culprit vessels. Patients underwent less revascularization in the FFR-guided group at 12 months [79.0 versus 86.8%, difference 7.8% (20.2%, 15.8%), P=0.054]^10^. In the FLOWER-MI trial, patients with ST-elevation myocardial infarction (STEMI) that underwent complete revascularization, the FFR guided group showed no significant benefit over an angiography, regarding the risk of death, MI, or urgent revascularization at 1 year for non-culprit vessels^14^.

In addition, FFR versus angiographic evaluation to guide CABG showed no impact on graft patency rate in two small-sized randomized clinical trials. In the FARGO trial, at 6 months, for 72 patients in the cohort, similar graft failure rates (16% versus 12%; p = 0.97) and clinical outcomes (rates of death, MI, and stroke) were seen in both the FFR and angiography guided groups^25^. Similarly in the GRAFFITI trial, no difference was seen in graft patency at one year^13^.

FAME 3 trial was one of the largest clinical trials comparing the noninferiority of FFR guided PCI using the latest-generation stents to conventional angiography guided CABG for patients with three-vessel disease. The results showed no difference in clinical outcomes at 1 year between the two groups^18^.

Prior meta-analyses investigating clinical outcomes in FFR versus angiography-guided PCI are outdated and lack current evidence. Furthermore, these analyses include non-randomized and observational studies which confer more bias than randomized control trials^26^. Our meta-analysis includes a comprehensive overview of the role of FFR versus angiography-guided revascularization in both patients with stable and unstable CAD as studied in randomized control trials. Also, we included patients undergoing FFR guided CABG. A recent meta-analysis investigated the role of FFR versus angiography to guide revascularization in patients with STEMI and multivessel disease for non-culprit lesions. However, patients were not randomized to FFR or angiography to guide PCI (apart from in FLOWER-MI), and thus there is no direct comparison between angiography and FFR-guided revascularization of non-culprit lesions^27^.

Our meta-analysis reflects the real-world population with a mix of patients with stable CAD, ACS, single vessel CAD as well as multivessel disease who are deemed candidates for revascularization with PCI and/or CABG. Our analysis shows that FFR provides no additional clinical benefit over conventional angiography. However, FFR can help reduce the number of stents used, prevent unnecessary revascularization procedures and is therefore more cost-effective when compared with angiography^28^.

## LIMITATIONS

There are several limitations in our study. First, there was no patient-level data available for our analysis. Additionally, there is heterogeneity in study designs, some studies compared FFR versus angiography guided PCI whereas other studies compared FFR versus angiography guided revascularization (PCI and CABG). However, sensitivity analysis was similar across all groups. FFR cutoff values also varied between studies and ranged from less than 0.75 to less than or equal to 0.80. Specifically, the RIPCORD study evaluated lesions <50% which maybe stretching the limit of FFR as there is no expected benefit of its use in these small lesions. The significant angiographic stenosis was based on operator discretion and ranged between 30% and 70%. Definitions of MACE were variable between studies. The duration of follow-up time also differed amongst the studies. Finally, none of the studies looked at patients exclusively with stable CAD.

## CONCLUSION

There was no difference in clinical outcomes in patients undergoing FFR-guided versus angiography-guided revascularization for patients with CAD.

## Data Availability

The data that support the findings of this study are available from the corresponding author upon reasonable request.

## ACKNOWLEDGEMENT, SOURCES OF FUNDING & DISCLOSURES

a) **Acknowledgments: None**
b) **Funding/Support:** None
c) **Conflict of Interest Disclosures:** No disclosures for the manuscript

## Supplemental Materials

Tables S1 Figures S1-S4

## Registration information

This meta-analysis was registered at PROSPERO (CRD42022301144). A review protocol was not prepared.

## REFERENCES

1. Fischer JJ, Samady H, McPherson JA, et al. Comparison between visual assessment and quantitative angiography versus fractional flow reserve for native coronary narrowings of moderate severity. Am J Cardiol. 2002;90(3):210–215. doi:10.1016/S0002-9149(02)02456-6

2. Kern MJ, Lerman A, Bech JW, et al. Physiological assessment of coronary artery disease in the cardiac catheterization laboratory: A scientific statement from the American Heart Association Committee on diagnostic and interventional cardiac catheterization, council on clinical cardiology. Circulation. 2006;114(12):1321–1341. doi:10.1161/CIRCULATIONAHA.106.177276

3. Tonino P Al, De Bruyne B, Pijls NH, et al. Fractional Flow Reserve versus Angiography for Guiding Percutaneous Coronary Intervention. Vol 360.; 2009.

4. Pijls NHJ, Fearon WF, Tonino PAL, et al. Fractional flow reserve versus angiography for guiding percutaneous coronary intervention in patients with multivessel coronary artery disease: 2-Year follow-up of the FAME (fractional flow reserve versus angiography for multivessel evaluation) study. J Am Coll Cardiol. 2010;56(3):177–184. doi:10.1016/j.jacc.2010.04.012

5. Chowdhury M, Osborn EA. Physiological Assessment of Coronary Lesions in 2020. Curr Treat Options Cardiovasc Med. 2020;22(1):2. doi:10.1007/S11936-020-0803-7

6. Pijls NHJ, Van Gelder B, Van Der Voort P, et al. Fractional Flow Reserve. Circulation. 1995;92(11):3183–3193. doi:10.1161/01.CIR.92.11.3183

7. Pijls NHJ, de Bruyne B, Peels K, et al. Measurement of fractional flow reserve to assess the functional severity of coronary-artery stenoses. N Engl J Med. 1996;334(26):1703–1708. doi:10.1056/NEJM199606273342604

8. Pijls NHJ, Van Gelder B, Van Der Voort P, et al. Fractional flow reserve. A useful index to evaluate the influence of an epicardial coronary stenosis on myocardial blood flow. Circulation. 1995;92(11):3183–3193. doi:10.1161/01.CIR.92.11.3183

9. Tonino PAL, Fearon WF, De Bruyne B, et al. Angiographic versus functional severity of coronary artery stenoses in the FAME study fractional flow reserve versus angiography in multivessel evaluation. J Am Coll Cardiol. 2010;55(25):2816–2821. doi:10.1016/J.JACC.2009.11.096

10. Layland J, Oldroyd KG, Curzen N, et al. Fractional flow reserve vs. angiography in guiding management to optimize outcomes in non-ST-segment elevation myocardial infarction: the British Heart Foundation FAMOUS-NSTEMI randomized trial. Eur Heart J. 2015;36(2):100–111. doi:10.1093/EURHEARTJ/EHU338

11. Van Nunen LX, Zimmermann FM, Tonino PAL, et al. Fractional flow reserve versus angiography for guidance of PCI in patients with multivessel coronary artery disease (FAME): 5-year follow-up of a randomised controlled trial. Lancet. 2015;386(10006):1853–1860. doi:10.1016/S0140-6736(15)00057-4

12. Bech GJW, De Bruyne B, Pijls NHJ, et al. Fractional flow reserve to determine the appropriateness of angioplasty in moderate coronary stenosis: a randomized trial. Circulation. 2001;103(24):2928–2934. doi:10.1161/01.CIR.103.24.2928

13. Toth GG, de Bruyne B, Kala P, et al. Graft patency after FFR-guided versus angiography-guided coronary artery bypass grafting: The GRAFFITI trial. EuroIntervention. 2019;15(11):E999–E1005. doi:10.4244/EIJ-D-19-00463

14. Puymirat E, Cayla G, Simon T, et al. Multivessel PCI Guided by FFR or Angiography for Myocardial Infarction. N Engl J Med. 2021;385(4):297–308. doi:10.1056/nejmoa2104650

15. Rioufol G, Dérimay F, Roubille F, et al. Fractional Flow Reserve to Guide Treatment of Patients With Multivessel Coronary Artery Disease. J Am Coll Cardiol. 2021;78(19):1875–1885. doi:10.1016/j.jacc.2021.08.061

16. Stables RH, Mullen LJ, Elguindy M, et al. Routine Pressure Wire Assessment Versus Conventional Angiography in the Management of Patients With Coronary Artery Disease: The RIPCORD 2 Trial. Circulation. 2022;146(9):687–698. doi:10.1161/CIRCULATIONAHA.121.057793

17. Moher D, Liberati A, Tetzlaff J, Altman DG. Preferred reporting items for systematic reviews and meta-analyses: The PRISMA statement. Int J Surg. 2010;8(5):336–341. doi:10.1016/j.ijsu.2010.02.007

18. Fearon WF, Zimmermann FM, De Bruyne B, et al. Fractional Flow Reserve– Guided PCI as Compared with Coronary Bypass Surgery. N Engl J Med. Published online November 4, 2021. doi:10.1056/nejmoa2112299

19. Mehta SR, Wood DA, Storey RF, et al. Complete Revascularization with Multivessel PCI for Myocardial Infarction. N Engl J Med. 2019;381(15):1411–1421. doi:10.1056/NEJMOA1907775/SUPPL_FILE/NEJMOA1907775_DATA-SHARING.PDF

20. Gershlick AH, Khan JN, Kelly DJ, et al. Randomized Trial of Complete Versus Lesion-Only Revascularization in Patients Undergoing Primary Percutaneous Coronary Intervention for STEMI and Multivessel Disease: The CvLPRIT Trial. J Am Coll Cardiol. 2015;65(10):963–972. doi:10.1016/J.JACC.2014.12.038

21. Lee JM, Kim HK, Park KH, et al. Fractional flow reserve versus angiography-guided strategy in acute myocardial infarction with multivessel disease: a randomized trial. Eur Heart J. Published online December 20, 2022. doi:10.1093/EURHEARTJ/EHAC763

22. Zhang Z, Li K, Tian J. Efficacy and safety outcomes of fractional flow reserve in guiding clinical therapy of non-ST-segment elevation myocardial infarction compared with angiography alone in elderly Chinese patients. Clin Interv Aging. 2016;11:1751–1754. doi:10.2147/CIA.S123735

23. Quintella EF, Ferreira E, Azevedo VMP, et al. Clinical outcomes and cost-effectiveness analysis of FFR compared with angiography in multivessel disease patient. Arq Bras Cardiol. 2019;112(1):40–47. doi:10.5935/abc.20180262

24. Park SH, Jeon KH, Lee JM, et al. Long-Term Clinical Outcomes of Fractional Flow Reserve-Guided Versus Routine Drug-Eluting Stent Implantation in Patients with Intermediate Coronary Stenosis: Five-Year Clinical Outcomes of DEFER-DES Trial. Circ Cardiovasc Interv. 2015;8(12). doi:10.1161/CIRCINTERVENTIONS.115.002442

25. Thuesen AL, Riber LP, Veien KT, et al. Fractional Flow Reserve Versus Angiographically-Guided Coronary Artery Bypass Grafting. J Am Coll Cardiol. 2018;72(22):2732–2743. doi:10.1016/J.JACC.2018.09.043

26. Enezate T, Omran J, Al-Dadah AS, et al. Fractional flow reserve versus angiography guided percutaneous coronary intervention: An updated systematic review. Catheter Cardiovasc Interv. 2018;92(1):18–27. doi:10.1002/CCD.27302

27. Bainey KR, Engstrøm T, Smits PC, et al. Complete vs Culprit-Lesion-Only Revascularization for ST-Segment Elevation Myocardial Infarction: A Systematic Review and Meta-analysis. JAMA Cardiol. 2020;5(8):1. doi:10.1001/JAMACARDIO.2020.1251

28. Hong D, Kim H, Lee H, et al. Long-Term Cost-Effectiveness of Fractional Flow Reserve–Based Percutaneous Coronary Intervention in Stable and Unstable Angina. JACC Adv. 2022;1(5):100145. doi:10.1016/j.jacadv.2022.100145

